# From Text to Tables: A Local Privacy Preserving Large Language Model for Structured Information Retrieval from Medical Documents

**DOI:** 10.1101/2023.12.07.23299648

**Authors:** Isabella C. Wiest, Dyke Ferber, Jiefu Zhu, Marko van Treeck, Sonja K. Meyer, Radhika Juglan, Zunamys I. Carrero, Daniel Paech, Jens Kleesiek, Matthias P. Ebert, Daniel Truhn, Jakob Nikolas Kather

**Author notes:** Corresponding author address: Jakob Nikolas Kather, MD, MSc Professor of Clinical Artificial Intelligence Else Kröner Fresenius Center for Digital Health Technische Universität Dresden DE – 01062 Dresden Mail ORCID ID: 0000-0002-3730-5348. **Disclosures** Jakob Nikolas Kather declares consulting services for Owkin, France; DoMore Diagnostics, Norway; Panakeia, UK; Scailyte, Basel, Switzerland; and Mindpeak, Hamburg, Germany; furthermore JNK holds shares in Kather Consulting, Dresden, Germany; and StratifAI GmbH, Dresden, Germany, and has received honoraria for lectures and advisory board participation by AstraZeneca, Bayer, Eisai, MSD, BMS, Roche, Pfizer and Fresenius. Daniel Truhn has received honoraria for lectures for Bayer and holds shares in StratifAI GmbH, Dresden, Germany. The authors have no other financial or non-financial conflicts of interest to disclose.

## Abstract

**Background and Aims:** Most clinical information is encoded as text, but extracting quantitative information from text is challenging. Large Language Models (LLMs) have emerged as powerful tools for natural language processing and can parse clinical text. However, many LLMs including ChatGPT reside in remote data centers, which disqualifies them from processing personal healthcare data. We present an open-source pipeline using the local LLM “Llama 2” for extracting quantitative information from clinical text and evaluate its use to detect clinical features of decompensated liver cirrhosis.

**Methods:** We tasked the LLM to identify five key clinical features of decompensated liver cirrhosis in a zero- and one-shot way without any model training. Our specific objective was to identify abdominal pain, shortness of breath, confusion, liver cirrhosis, and ascites from 500 patient medical histories from the MIMIC IV dataset. We compared LLMs with three different sizes and a variety of pre-specified prompt engineering approaches. Model predictions were compared against the ground truth provided by the consent of three blinded medical experts.

**Results:** Our open-source pipeline yielded in highly accurate extraction of quantitative features from medical free text. Clinical features which were explicitly mentioned in the source text, such as liver cirrhosis and ascites, were detected with a sensitivity of 100% and 95% and a specificity of 96% and 95%, respectively from the 70 billion parameter model. Other clinical features, which are often paraphrased in a variety of ways, such as the presence of confusion, were detected only with a sensitivity of 76% and a specificity of 94%. Abdominal pain was detected with a sensitivity of 84% and a specificity of 97%. Shortness of breath was detected with a sensitivity of 87% and a specificity of 96%. The larger version of Llama 2 with 70b parameters outperformed the smaller version with 7b parameters in all tasks. Prompt engineering improved zero-shot performance, particularly for smaller model sizes.

**Conclusion:** Our study successfully demonstrates the capability of using locally deployed LLMs to extract clinical information from free text. The hardware requirements are so low that not only on-premise, but also point-of-care deployment of LLMs are possible.

**Lay summary:** We leveraged the large language model Llama 2 to extract five key features of decompensated liver cirrhosis from medical history texts, simplifying the analysis of complex text-based healthcare data.

## Introduction

It is estimated that 80% of clinical data exists in an unstructured format.^1^ This “dark matter” of healthcare data is currently unusable for quantitative computational analysis. While deep learning methods have made structured data from Electronic Health Records (EHRs) usable for individual risk prediction,^2^ and can make diagnoses and extract biomarkers from radiology or histopathology images,^3,4^ natural language has not been widely used as a source to extract structured information. Making an unstructured data resource readable for downstream tasks has a variety of benefits, such as improvements in individual healthcare outcomes,^5^ the possibility to obtain scientific insight,^6^ and improvement in billing processes and quality control.^7^

In Natural Language Processing (NLP) computational methods are applied to unstructured text. Medical applications of NLP have been explored for decades,^8,9^ but real-world applications are still very rare. However, real world data analysis becomes increasingly acknowledged and implemented in timely evidence generation which makes the need to extract real world data from text even more pressing.^10^ Several hurdles have been discussed for NLP in healthcare, among them the lack of annotated datasets and user-centered design as well as hand-crafted over-engineered software pipelines which lack scalability.^11,12^ LLMs have changed this field: they are transformer neural networks which are trained on large bodies of unstructured text data with self-supervised learning (SSL).^13–16^ LLMs are foundation models which can be applied to a broad range of tasks without having been explicitly trained for these tasks. This “zero-shot” application changes the conventional wisdom in medical artificial intelligence by which a model for a certain task needs to be trained on a large dataset representing this specific task.^17^ In particular, the LLM Generative Pretrained Transformer (GPT) and its user interface ChatGPT, have demonstrated remarkable proficiency in structuring text and extracting relevant information in a quantitative way.^18^ Their capabilities could revolutionize the way we comprehend and process vast quantities of healthcare data.^19–21^ For example, GPT-4 has been used to extract structured clinical information from free text reports in radiology,^18^ pathology and medicine.^22^

However, these LLMs run as cloud services and using them requires the transfer of privileged information to remote servers. This creates massive legal and ethical challenges, especially in the European Union (EU), where the export of personal health data is not possible.^23,24^ Ideally, LLMs should run on premise of healthcare institutions, potentially even at the point of care.^25,26^ However, this requires software pipelines using lightweight LLMs, which are currently not validated for medical tasks. Here, we aimed to build and validate a fully automated pipeline for end-to-end processing of clinical text data which uses locally deployable LLMs and can potentially be used at the point of care. We investigated the capabilities of our new pipeline on a task of high clinical importance, the identification of decompensated liver cirrhosis. Approximately 1% of the population in the EU has liver cirrhosis^27^ and decompensation is among the most common emergencies in these patients.^28^ Decompensation is often overlooked initially, but can be a turning point in the prognosis of cirrhotic patients, thus early identification and management are crucial to improve patient outcomes.^29^ Having an automated detection of liver decompensation from clinical free text could be the basis of early warning systems in clinical routine. Furthermore, this could facilitate retrospective analysis of clinical data for scientific, quality control or billing purposes.

## Methods

### Ethics statement

We solely utilized anonymized patient data from the MIMIC IV database. The MIMIC IV dataset is a comprehensive and publicly available collection of anonymized medical data from patients admitted to the emergency department or intensive care unit at Beth Israel Deaconess Medical Center in Boston Massachusetts, United States and enables text based research in healthcare and serves as a benchmark for medical AI studies.^30^ The MIMIC IV database contains a broad spectrum of patient data collected from 2009 to 2019, thereby being representative of multiple clinical scenarios.^31^ All research procedures were conducted in accordance with the Declaration of Helsinki.

### Data preparation

We applied for access to the MIMIC-IV database available from physionet.org and obtained access to the comprehensive health-related data of patients treated in an emergency department or intensive care setting.^30^ Central to our study was the early detection of decompensated liver cirrhosis in admission records, a critical task due to the condition’s potential lethality and rapid progression to complications such as variceal bleeding, hepatic encephalopathy, or renal failure. Early and accurate identification is vital for initiating immediate treatment and guiding patient management. For this study, we selected the first 500 patient histories (0,15% of all MIMIC IV clinical notes), focusing on identifying signs of decompensation in liver cirrhosis. We utilized Llama 2 to extract three symptoms - shortness of breath, abdominal pain, and confusion - from the text, and to identify two explicitly stated conditions: liver cirrhosis and ascites. This approach aimed to demonstrate the model’s effectiveness in discerning both implicit and explicit medical information crucial for patient care.

### Model details and data processing

The study’s goal was to assess the capability of the LLM “Llama 2”, in extracting the mentioned information from the textual medical data. We employed the zero-shot method to run the model. In our approach, all three versions of Llama 2 were used, the 7 billion-, 13 billion-, and 70 billion parameter-sized model. Our aim was to retrieve information about the five predefined features from patients’ present medical histories.^30^ Initially, the model was prompted to give JSON formatted output, but the model’s JSON output was inconsistent and defective. Therefore, we utilized the llama.cpp version,^32^ a framework originally designed for running Llama 2 models on lower-resource hardware and beyond that supporting grammar-based output formatting. Thus, we enforced JavaScript Object Notation (JSON) format generation using llama.cpp’s grammar-based sampling, which dictates text generation through specific grammatical rules to ensure valid JSON. We then converted these JSON outputs into CSV format using Python’s pandas library. The whole pipeline is depicted in **Figure 1**.

**Figure 1.**
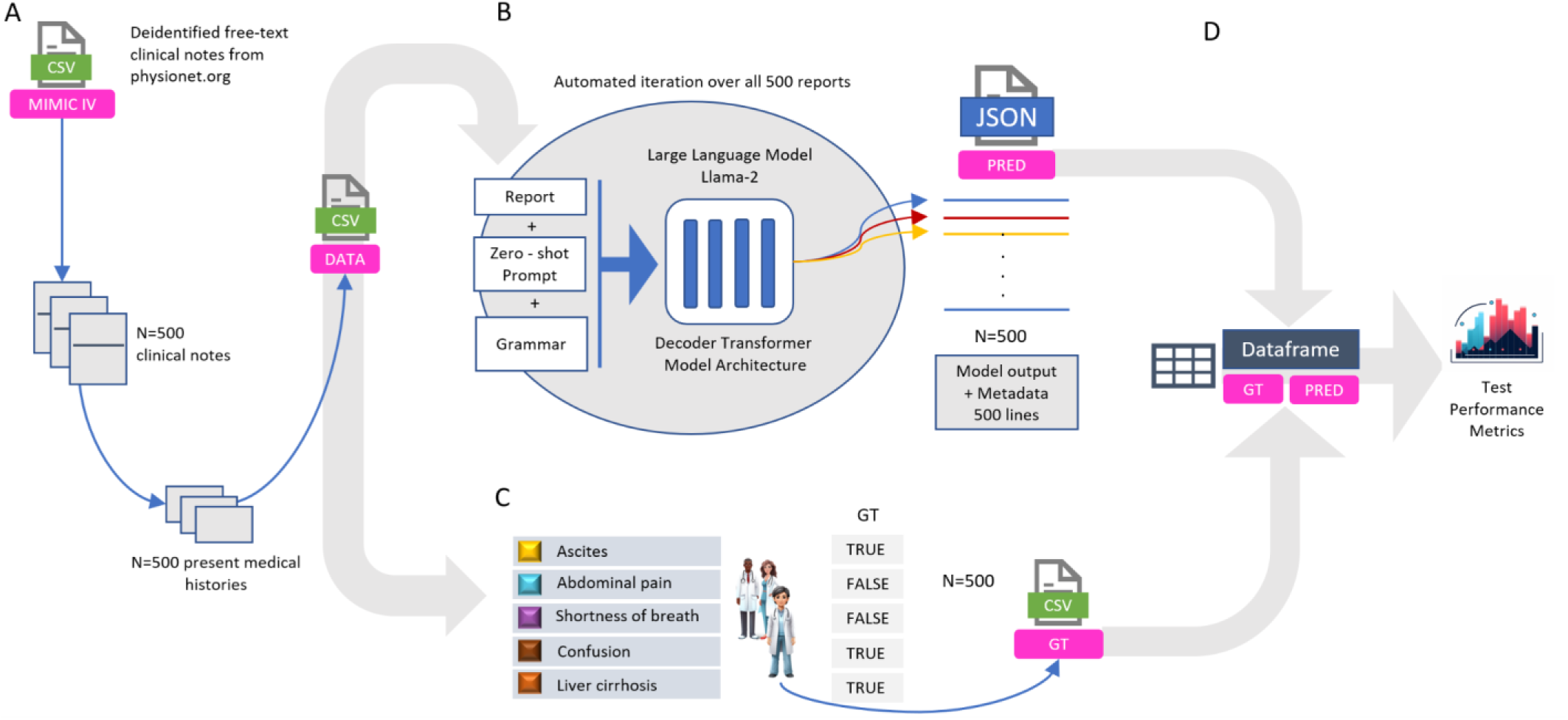
Experimental design and Feature Extraction Pipeline. **A** We implemented an automated process to extract 500 free-text clinical notes from the MIMIC IV database, focusing specifically on the patients’ present medical histories. These selected anamnesis reports were then systematically converted and stored in a CSV file for further processing. **B** Utilizing this CSV file, our custom-designed software algorithm selected one report at a time and combined it with a predetermined prompt and grammatical structures. This combination was then input into the advanced large language model, Llama 2. The primary function of Llama 2 in our study was to meticulously identify and extract specific, predefined clinical features (namely, Shortness of Breath, Abdominal Pain, Confusion, Ascites, and Liver Cirrhosis) from the clinical reports. The extracted data was subsequently formatted into a JSON file. To ensure a high degree of precision and structured output, we applied a grammar-based sampling technique. **C** To establish a benchmark, we engaged three medical experts who independently analyzed the same clinical reports. They extracted identical items as the Llama 2 model, thereby creating a reliable ‘ground truth’ dataset. **D** This ground truth dataset served as a reference point for a quantitative comparison and analysis of the model’s performance, assessing the accuracy and reliability of the information extracted by Llama 2. Icon source: Midjourney ^43^

### Prompt engineering

We implemented a technique known as zero-shot chain-of-thought prompting, wherein the model is tasked with identifying relevant text passages without prior training specific to the task—this is the “zero-shot” aspect, which tests the model’s ability to apply its pre-trained knowledge to new problems. To add explainability to the models’ answers, we forced the model to use a certain style when providing its answer. This also implented a “chain-of-thought” process, which allowed sequential reasoning where the LLM output transparently outlines its thought process, to verify the existence of a particular feature within the text. To enhance outcomes via prompt engineering, one-shot prompting was also employed^33^, providing the model with an example report and corresponding JSON formatted output. Blinded medical raters established a consensus on precise definitions for the queried features during ground truth definition, which were subsequently provided to the model (definition prompting). Ultimately, single-shot and definition chain-of-thought prompting were combined. The standard Llama 2 prompt contains two modules, the “system” and the “user” part. The system prompt provides initial instructions or explanations to guide the interaction, while the user prompt includes the user’s input or query, further shaping the response process. We experimented with different arrangements of system and user prompts in combination with definition, one-shot and chain-of-thought prompting. Our detailed prompt engineering approach is described in the Supplementary methods.

### Ground Truth definition

For validation, the 500 reports were meticulously and independently assessed by clinicians to establish a ground truth. In the event of disagreement, a consensus was always reached through discussion. A comprehensive overview regarding consensus about the ground truth rating, as well as challenges and methodologies concerning ground truth definition, can be found in the Appendix.

### Evaluation of model results

Positive Predictive Value (Precision, PPV), Sensitivity (Recall), Specificity, Negative Predictive Value (NPV) and Accuracy were computed to assess the performance of the different model’s outputs. To obtain reliable estimates, we employed bootstrapping, a statistical resampling technique, executing 1000 iterations. This method involves repeatedly sampling from the dataset with replacement to create many “bootstrap” samples. These samples are then used to estimate the variability and confidence of our statistical estimates, enhancing their robustness and credibility. All source codes are available at https://github.com/I2C9W/fromtexttotables/releases/tag/v0.5.0.

## Results

### Key Medical Features are unevenly represented in Medical Histories

Our analysis of the Llama 2 model’s data extraction capabilities from text reports focused on five key medical features: liver cirrhosis, ascites, abdominal pain, shortness of breath, and confusion. We found that the frequency of these features varied significantly across the reports. Abdominal pain and shortness of breath were frequently documented in the data. However, liver cirrhosis and ascites were less prevalent, mentioned in only about 5% of cases, as detailed in **figure 2**.

**Figure 2.**
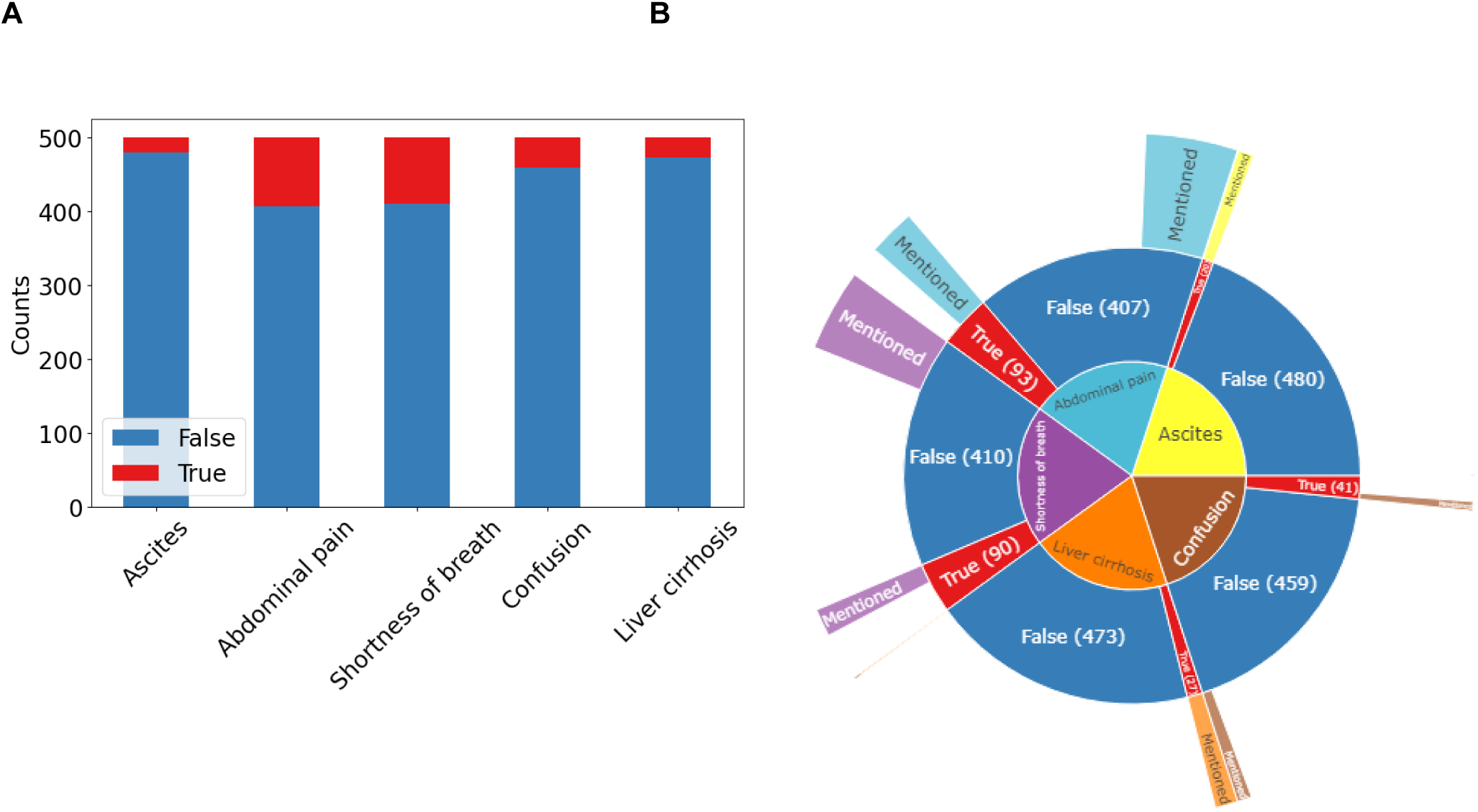
Feature Distribution in 500 MIMIC present medical histories. **A** The bar chart visualizes data from 500 present medical history reports extracted from the MIMIC-IV database. It displays the counts for five extracted features, with ‘true’ counts in red and ‘false’ in blue. **B** The sunburst plot indicates the amount of reports, in which the features’ term is explicitly mentioned as a share of false and true counts. Liver cirrhosis and ascites are the most frequently explicitly mentioned features, with every mention aligning with a ‘true’ classification in the ground truth evaluation. Abdominal pain and shortness of breath were most frequently mentioned over all reports.

While liver cirrhosis and ascites were explicitly mentioned when present (ascites was mentioned in 19 reports and also present in 19 reports), making their detection more straightforward, the documentation of abdominal pain, shortness of breath, and confusion often required more nuanced interpretation, as these symptoms were described in multiple ways by physicians. These symptoms were not always explicitly stated but could be inferred or deduced from contextual information. For example, abdominal pain might be indicated through a variety of descriptors or understood from the absence of certain findings, e.g. “pain in the RUQ” stands for “pain of the right upper quadrant of the abdomen” thus indicating the presence of abdominal pain.

Similarly, shortness of breath and confusion, while not always directly stated, could be inferred from contextual clues or specific medical terminology used in the reports. This implies that accurately identifying such implicit features demands a nuanced understanding of medical language and context, as well as some level of clinical expertise. For example, a statement like “10-point review of systems negative” implies the absence of symptoms like shortness of breath, abdominal pain, and confusion, requiring the model to interpret these indirect cues effectively.

### Llama 2 is able to extract relevant information from unstructured text

In our assessment, the 70b model displayed remarkable proficiency. Sensitivity of detecting liver cirrhosis and ascites was 100% and 95%, respectively. For abdominal pain and shortness of breath, sensitivities were lower with 84% and 87%, respectively. Confusion was the symptom which was most difficult to extract for the LLM with a sensitivity of only 76%. Specificity for liver cirrhosis was 96%, for ascites 95% and even higher for abdominal pain (97%), shortness of breath (96%) and confusion (94%). Confusion matrices are shown in **figure 3**.

**Figure 3.**
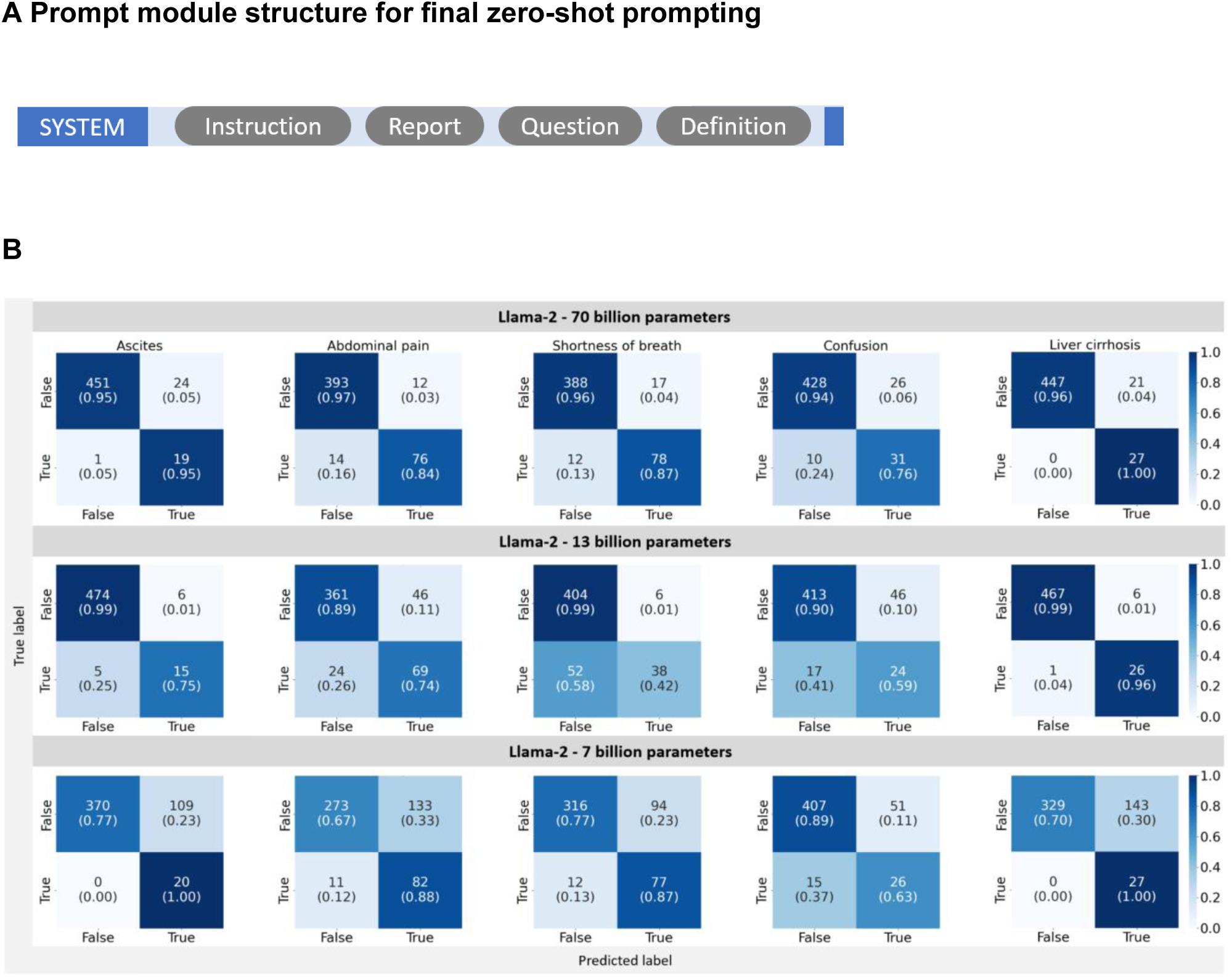
Confusion matrices for extracted features with zero-shot prompting. The confusion matrices visualize the performance of the Llama 2 models with 7 billion, 13 billion and 70 billion parameters in retrieving the presence or absence of the five features ascites, abdominal pain, shortness of breath, confusion and liver cirrhosis in all n=500 medical histories from MIMIC IV. All matrices are divided into four quadrants with the two labels “true” or “false” in each axis. The x-axis depicts the predicted labels, the y-axis depicts the true labels. The confusion matrices are normalized to show proportions, where each cell represents the fraction of predictions within the actual class. Values along the diagonal indicate correct predictions (true positives and true negatives), while off-diagonal values represent misclassifications (false positives and false negatives). The sum of each row’s fractions equals 1, indicating the proportion of predictions for each actual class. The ‘n’ values represent the absolute number of observations in each category. In the bottom left matrix, the extraction of ascites with the 70b model is shown. The top left quadrant (true positives) shows a high score of 0.95, indicating a high rate of correct predictions for actual cases of Ascites. The top right quadrant (false negatives) has a score of 0.05, suggesting few cases were incorrectly predicted as not having Ascites. The bottom left quadrant (false positives) has a score of 0.1, indicating few cases were incorrectly identified as Ascites. Finally, the bottom right quadrant (true negatives) shows a high score of 0.9, which means a high rate of correct predictions for non-cases.

One-shot prompting yielded slightly better results with higher sensitivities (ascites: 95%, abdominal pain: 92%, shortness of breath: 83%, confusion: 88% and liver cirrhosis 100%) and specificities (ascites: 99%, abdominal pain:92%, shortness of breath: 96%, confusion: 94% and liver cirrhosis 97%) (**Figure 4 and Supplementary Table 2**).

**Figure 4.**
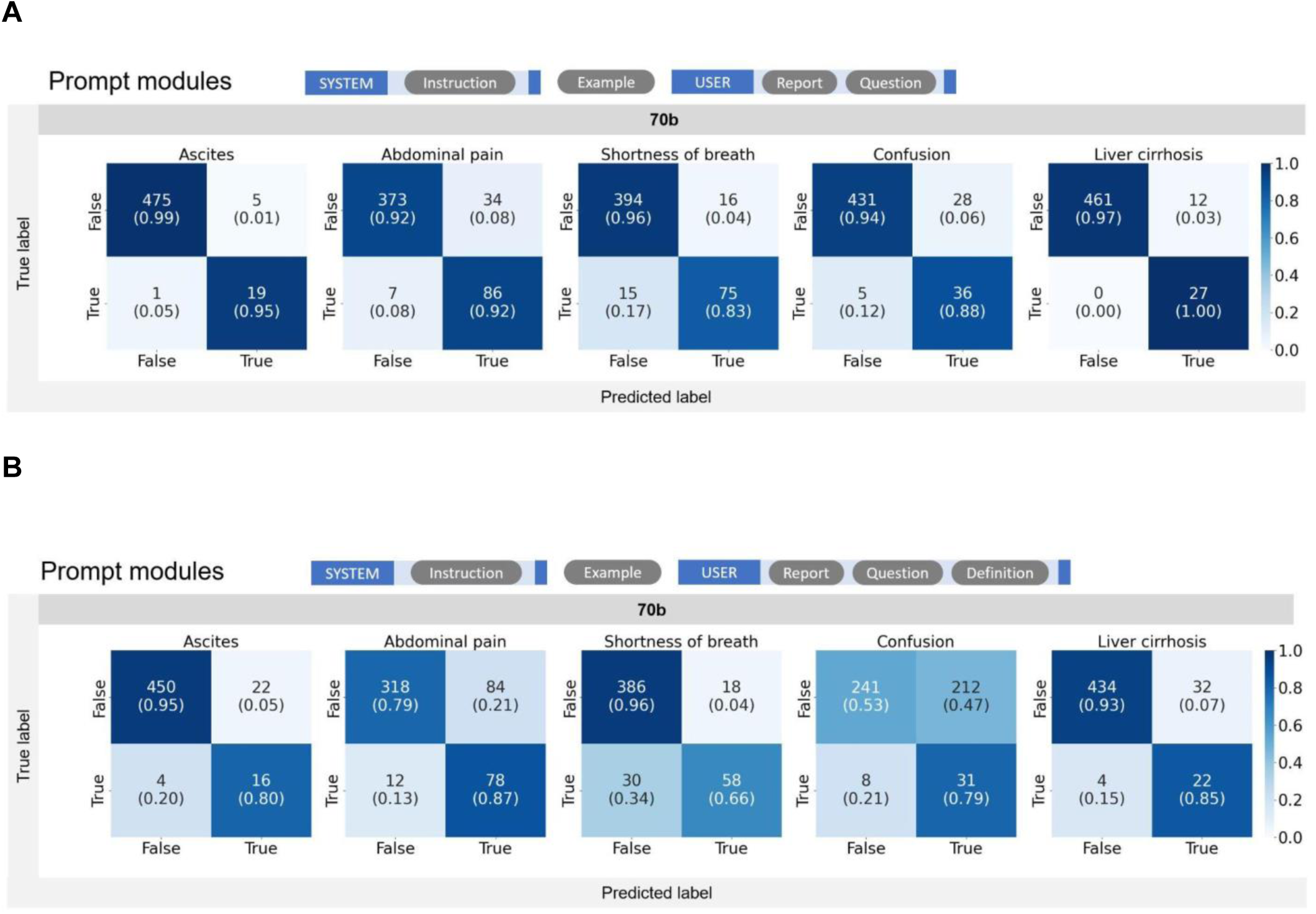
Confusion matrices for extracted features with one-shot prompting. The confusion matrices visualize the performance of the Llama 2 models with 70 billion parameters in retrieving the presence or absence of the five features ascites, abdominal pain, shortness of breath, confusion and liver cirrhosis in all n=500 medical histories from MIMIC IV. All matrices are divided into four quadrants with the two labels “true” or “false” in each axis. The x-axis depicts the predicted labels, the y-axis depicts the true labels. The confusion matrices are normalized to show proportions, where each cell represents the fraction of predictions within the actual class. Values along the diagonal indicate correct predictions (true positives and true negatives), while off-diagonal values represent misclassifications (false positives and false negatives). The numbers indicate absolute counts, the figure in brackets indicate fractions. The sum of each row’s fractions equals 1, indicating the proportion of predictions for each actual class. **A** shows the best one-shot prompt architecture and results. Whereas adding definitions, which improved performance with zero-shot prompting, deteriorated the results for one-shot prompting **(B)**.

The models with more parameters performed better, with the most substantial increase in accuracy from the Llama 2 7b to 13b model **(Supplementary Table 1, Figure 5)**. For implicit features, the 70b model yielded the highest accuracy. The 7b model faced challenges in accurately identifying false classifications. All models presented a high negative predictive value. Precision and specificity tended to improve most from 7b to 13b parameter model size. Recall was best in the explicitly mentioned features **(Supplementary Table 1 and Supplementary Table 2)**.

**Figure 5.**
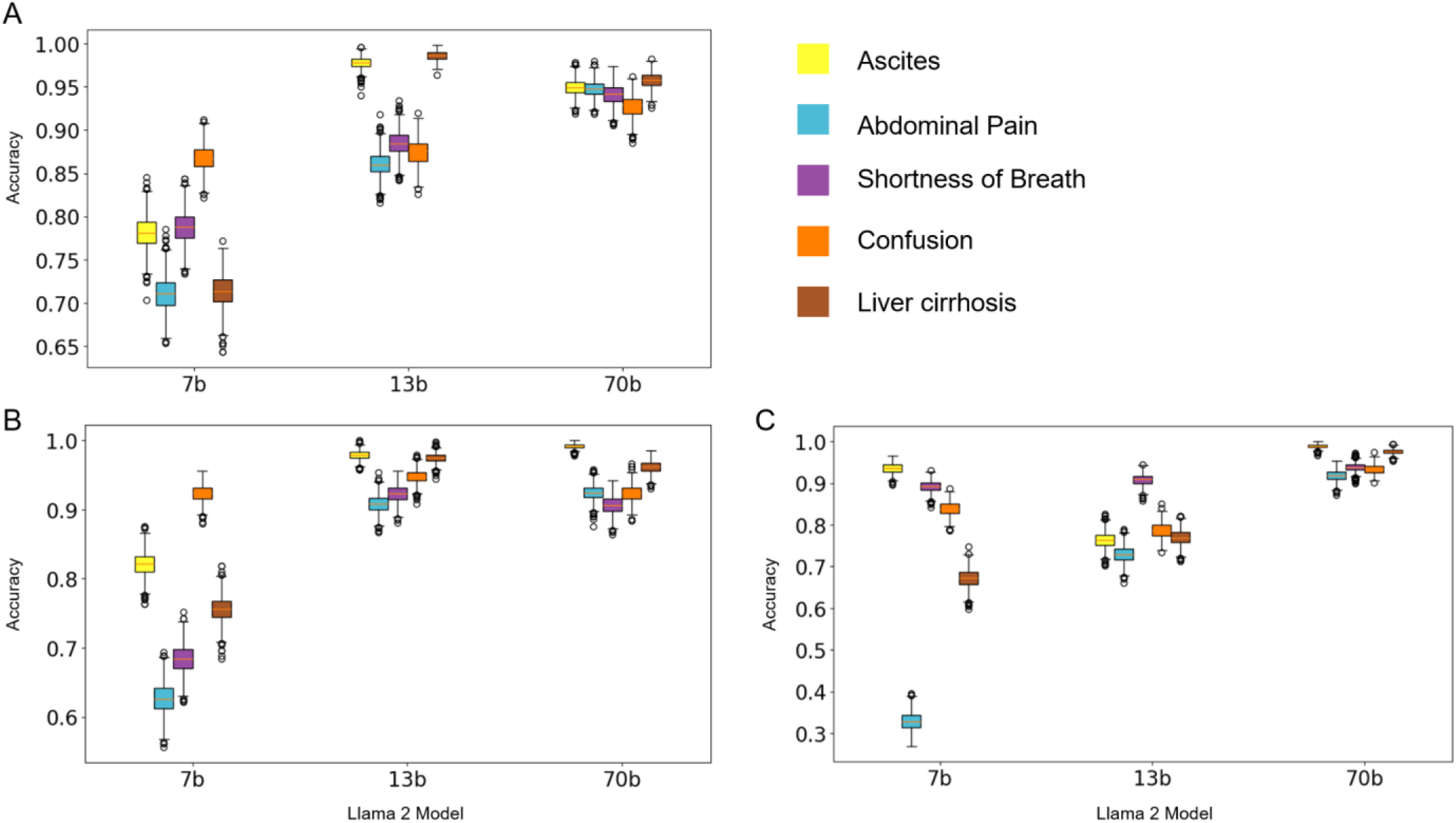
Accuracy for prediction of present features with different parameter size models. This graph compares the accuracy of different models (7b, 13b, and 70b) in extracting the five features Ascites, Abdominal pain, Shortness of breath, Confusion, Liver cirrhosis. **A** depicts the accuracy of the final zero-shot prompting, **B** with plain zero shot prompting without additional definition or example, **C** the accuracy of the best one-shot prompting example. Error bars represent the variability or confidence intervals, calculated with 1000-fold bootstrapping.

### Prompt engineering enhances accuracy, especially in smaller sized models

In our initial test, we tested with the 7b model and used a combination of a system prompt with general instructions and a user prompt containing the report and questions (prompting strategy details in **Supplementary Figure 2 and 3**). Including a one-shot example in the prompt slightly enhanced the model’s accuracy except for the feature abdominal pain (**Supplementary Figure 2**). The human instructions in the Llama prompt need to be indicated within specific tags ([INST],[/INST]). Notably, the one-shot example needed to be excluded from the instruction section, otherwise the performance deteriorated substantially, because the model answered the questions with the example given. Requesting an excerpt from the text followed by a binary answer (Chain-of-thought prompting) did not yield improved results, only for the feature liver cirrhosis. For explainability reasons, we nevertheless forced the model with the grammar (github repository) to provide an excerpt first and then the binary outcome and found that this did not adversely affect performance.

Providing definitions for all features only improved the extraction of the more implicitly mentioned features shortness of breath and abdominal pain, but deteriorated the extraction of explicitly mentioned features. Subsequent testing involved consolidating both the report and question components within the system prompt, instead of dividing them between system and user prompts. This change resulted in improved performance for the 7b model, whereas this trend was not consistently present for the 70b model (e.g. accuracy ascites (7b) 78% vs.82%, liver cirrhosis (7b) 69% vs. 76%, abdominal pain (7b) 57% vs. 63%, shortness of breath (7b) 64% vs. 68%, confusion 90% vs. 92%), indicating a more effective prompt structure when integrated into the system prompt (Supplementary Methods). Finally, the most effective prompt structure for zero-shot prompting, as concluded from our experiments, was to include all components within the system prompt. This encompassed providing a report, asking specific questions, giving definitions for implicit features, and enforcing a chain-of-thought response through grammatical structuring without a chain-of-thought questioning strategy. Nevertheless, the prompt experiments changed each feature differently. In principle, the least differences between the prompting techniques can be seen in the largest, 70b model. In summary, these data show that prompt engineering can help improving performance especially in the smallest model, whereas larger model sizes demonstrated greater robustness, with remarkably high performance of simple prompts, improving only marginally through prompt engineering.

## Discussion

In this study, we present an open-source software pipeline which can use local LLMs to extract quantitative data from clinical free text and evaluate it on the detection of symptoms indicating decompensated liver cirrhosis, an important medical emergency. We demonstrate that the lightweight LLM “Llama 2” yields an excellent performance on this task, even in a zero-shot way without any task-specific retraining. Specifically, the 70 billion parameter model was able to achieve 90% accuracy or more for both implicitly and explicitly mentioned features. Historically, rule-based or dictionary-based methods were used for information extraction^34^, but these approaches struggle with the variability of medical texts and the scarcity of labeled training data.^35^ However, such rule-based hand-crafted methods cannot extract implicitly stated information in a zero-shot way. Therefore, we show that LLMs can fill the gap in information extraction and will be of utmost importance for versatile healthcare data processing.

The performance of LLMs is massively increasing month to month^36^ and we expect that future LLMs will further improve the performance. Many proof-of-concept studies for LLMs in medicine only show a semiquantitative analysis — in contrast, we employ a rigorous, quantitative, pre-specified analysis comparing the models’ outputs to a ground truth obtained by three blinded observers. We posit that such a systematic analysis should be the gold standard in assessing the benefits and shortcomings of LLMs in medicine.

Not surprisingly, we find that clinical features that are explicitly mentioned in clinical texts are recalled more effectively by our model than those that are implied, indicating a limited grasp of contextual subtleties. The model particularly struggled with extracting ‘confusion’ due to inconsistent documentation and definition, which even required medical experts to consent about a definition (see **Supplementary Tables 3 and 4** in the Appendix for raters’ agreement and feature consensus definition). Despite this, the Llama 2 70b model excels in identifying implicitly mentioned features, showing a superior understanding of context linked to its larger parameter size. Our prompt experiments’ findings indicate that models with larger parameter size demonstrate enhanced robustness, and their performance remains largely unaffected by variations in prompt engineering, suggesting promising prospects for the development of even better and larger models in the future. Llama has been previously successful in tasks like DRG prediction and ICD code extraction from clinical notes.^37,38^ Our analysis reaffirms Llama 2’s strong information extraction capabilities and secure processing of sensitive patient data. Nevertheless, Llama as a decoder-only model has proven to struggle more with unseen information types than encoder-decoder models,^39^ although decoder-only models with more extensive pre-training overcome this limitation. Continuous improvements to Llama and other large language models (LLMs), as seen with ChatGPT, could further boost their performance in complex tasks.^40^ Several related studies have shown that the LLM GPT-4 excels at structured information extraction from medical text and is often superior to Llama 2. However, GPT-4 runs in the cloud and its architecture is unknown to the public,^41^ making it currently not suitable for processing personal healthcare data.

LLMs have some fundamental limitations which users must be aware of. In our analysis, we encountered some of these: For instance, our analysis revealed that when Llama 2 was asked to determine a patient’s gender from medical history, it based its decision on the prevalence of certain symptoms in one gender over another, rather than using clear identifiers like personal pronouns, which prove the gender instead of suggesting it by probabilities (**Supplementary Figure 1**, Appendix). Addressing biases in LLMs is essential to ensure the accuracy and impartiality of the information they deliver. Continuous investigation and the development of advanced methods to assess these models’ functioning are vital. This will enable us to rely on these models for information that reflects the actual content, rather than assumptions made by the model. Furthermore, we analyzed Llama’s proficiency in evaluating english-language patient histories; its ability to handle data in other languages needs to be further elucidated, since 90% of Llama-2’s training data was English language data.^26^

Our analysis has the potential to form a basis for clinical decision support systems (CDSS), aiding in identifying symptoms of conditions like decompensated liver cirrhosis and applicable in various medical fields. Further refinement and evaluation, potentially through fine-tuning, retrieval augmented generation approaches^42^ and larger language models are necessary to obtain the necessary security in handling medical data. Nevertheless, our research reveals substantial chances for broader medical settings: Enhanced information extraction from free text enables more effective quantitative analysis in research. Moreover, it can streamline quality control in hospital procedures and simplify billing encoding, thereby reducing labor-intensive information extraction tasks.

## Data Availability

We used the MIMIC-IV dataset which is available after request from physionet.org.

https://github.com/I2C9W/fromtexttotables/releases/tag/v0.5.0

