## Appendix for "From Text to Tables: A Local Privacy Preserving Large Language Model for Structured Information Retrieval from Medical Documents"

**Supplementary Table 1 - Model Performance - Zero-shot prompting with definitions**

| | Sensitivity | | | Specificity | | | Positive predictive value | | | Negative predictive value | | | Accuracy | | |
|---|---|---|---|---|---|---|---|---|---|---|---|---|---|---|---|
| | **7b** | **13b** | **70b** | **7b** | **13b** | **70b** | **7b** | **13b** | **70b** | **7b** | **13b** | **70b** | **7b** | **13b** | **70b** |
| **Ascites** | 1.00 | 0.75 | 0.95 | 0.77 | 0.99 | 0.95 | 0.16 | 0.71 | 0.44 | 1.00 | 0.99 | 1.00 | 0.78 | 0.98 | 0.95 |
| **Abdominal Pain** | 0.88 | 0.74 | 0.84 | 0.67 | 0.89 | 0.97 | 0.38 | 0.60 | 0.86 | 0.96 | 0.94 | 0.97 | 0.71 | 0.86 | 0.95 |
| **Shortness of Breath** | 0.87 | 0.42 | 0.87 | 0.77 | 0.99 | 0.96 | 0.45 | 0.86 | 0.82 | 0.96 | 0.89 | 0.97 | 0.79 | 0.88 | 0.94 |
| **Confusion** | 0.63 | 0.59 | 0.76 | 0.89 | 0.90 | 0.94 | 0.34 | 0.34 | 0.54 | 0.96 | 0.96 | 0.98 | 0.87 | 0.87 | 0.93 |
| **Liver cirrhosis** | 1.00 | 0.96 | 1.00 | 0.70 | 0.99 | 0.96 | 0.16 | 0.81 | 0.56 | 1.00 | 1.00 | 1.00 | 0.71 | 0.99 | 0.96 |

Comparing three versions of Llama-v2, the largest (70b) models showed the highest performance whereas the smallest (7b) model performed worst. The 13b and 70b models show higher accuracy across all conditions when compared to the 7b model.

**Supplementary Table 2 - Model Performance - One-shot prompting**

| | Sensitivity | | | Specificity | | | Positive predictive value | | | Negative predictive value | | | Accuracy | | |
|---|---|---|---|---|---|---|---|---|---|---|---|---|---|---|---|
| | **7b** | **13b** | **70b** | **7b** | **13b** | **70b** | **7b** | **13b** | **70b** | **7b** | **13b** | **70b** | **7b** | **13b** | **70b** |
| **Ascites** | 0.95 | 1.00 | 0.95 | 0.94 | 0.76 | 0.99 | 0.38 | 0.13 | 0.79 | 1.00 | 1.00 | 1.00 | 0.94 | 0.76 | 0.99 |
| **Abdominal Pain** | 0.99 | 0.95 | 0.92 | 0.18 | 0.68 | 0.92 | 0.22 | 0.40 | 0.72 | 0.99 | 0.98 | 0.98 | 0.33 | 0.73 | 0.92 |
| **Shortness of Breath** | 0.64 | 0.59 | 0.83 | 0.95 | 0.98 | 0.96 | 0.72 | 0.87 | 0.82 | 0.92 | 0.91 | 0.96 | 0.89 | 0.91 | 0.94 |
| **Confusion** | 0.71 | 0.85 | 0.88 | 0.85 | 0.78 | 0.94 | 0.30 | 0.25 | 0.56 | 0.97 | 0.98 | 0.99 | 0.84 | 0.79 | 0.93 |
| **Liver cirrhosis** | 1.00 | 1.00 | 1.00 | 0.65 | 0.76 | 0.97 | 0.14 | 0.18 | 0.69 | 1.00 | 1.00 | 1.00 | 0.67 | 0.77 | 0.98 |

#### Supplementary Table 3 - Definition of extracted features for ground truth rating

| Variable | Definition |
|---|---|
| Shortness of breath | Any kind of dyspnoea, also dyspnoea on exertion (DOE) |
| Abdominal pain | Any kind of abdominal discomfort, including explicit pain, distention<br>Any kind of abdominal localisation (epigastric, all quadrants) |
| Confusion | Objectively stated altered mental status,<br>Subjectively reported altered mental status,<br>evaluated according to report (patient not oriented to time, location, person, situation)<br>10-point Review of Systems negative is equal to absent confusion |
| Ascites | Any kind of pathological accumulation of free fluid in the abdominal cavity |
| Liver cirrhosis | Chronic disease of the liver associated with destruction of lobular and vascular architecture by inflammatory fibrosis. |

**Supplementary Table 4 - Consensus for ground truth definition**

Medical documentation is often ambiguous. Information does not always correspond to the same level of precision, so agreement among raters was necessary to ensure a consistent definition of ground truth. Free-text medical documentation is often fuzzy, so mutually exclusive and commonly exhaustive categories cannot always be easily defined.

| | Shortness of breath | Abdominal pain | Confusion | Ascites | Liver cirrhosis |
|---|---|---|---|---|---|
| "10 point review of systems (ROS) negative" | negative | negative | negative | Not available | Not available |
| "No further complaints" | negative | negative | Not available | Not available | Not available |

"This is a ___ yo f with h/o recently diagnosed metastatic cancer of unknown prior presenting with nausea, vomiting, and fever to 101 today. (…) Pt. also reports pain over her RUQ and has very poor PO intake. Pt has been feeling progressively weak over this time period. (…) No URI symptoms, no urinary complaints. Pt has been constipated, which improves when stops her anti-emetics. Last bowel movement was yesterday. Pt is passing gas. Pt has lower extremity edema, which has been present for the past several weeks. Of note, Pt was supposed to have one of her liver mets biopsied in the past several weeks, but Pt was taking ibuprofen so the biopsy had to be postponed. In the ED, initial VS were: 97.6 117 128/74 18 95 RA. Labs were significant for WBC of 18.7, with 77% polys. UA was significant for ketones. Patient received zofran, NS. She had a CXR that showed new left sided opacity that may reflect PNA superimposed on metastatic disease vs. lymphangiitic spread of cancer. She received vanc and cefepime for pneumonia. ROS:(+) per HPI (-) night sweats, headache, vision changes, rhinorrhea, congestion, sore throat, BRBPR, melena, hematochezia, dysuria, hematuria."

**Prompt:**

What is the patient's gender?

**Answer Llama-2 7b:**

The patient's gender can be determined from their medical history. According to the information provided, the patient has recently been diagnosed with metastatic cancer of unknown origin, which suggests that they are likely female. Females are more susceptible to developing breast, ovarian, and uterine cancers, all of which are common sites of

**Supplementary Figure 1 - Example of biases of Large language models.** The figure displays a modified medical history report from MIMIC IV on the left side, in which the patient's gender is subtly indicated only by the abbreviation "f" and the personal pronoun "she," both highlighted in a red box. On the right side, when Llama 2 is prompted about the patient's gender, it fails to recognize these subtle indications. Instead, it infers the gender as female based on the likelihood of certain conditions mentioned in the report, rather than the explicit gender markers.

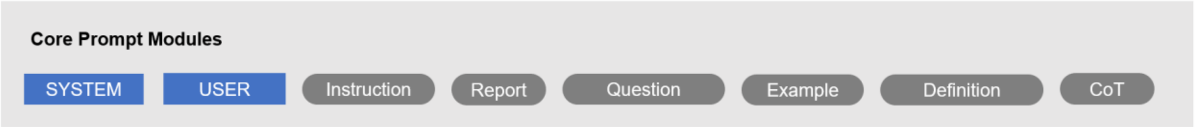


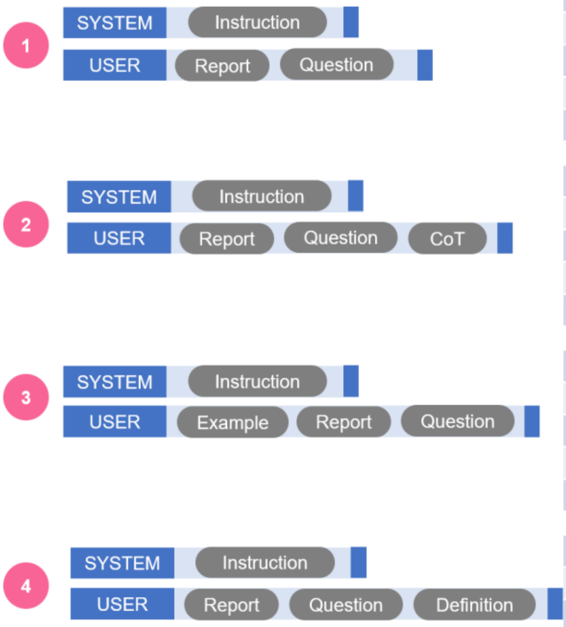


| Feature | Sensitivity | Specificity | Ppv | Npv | Accuracy |
|---|---|---|---|---|---|
| Ascites | 0.95 | 0.78 | 0.15 | 1.0 | 0.78 |
| Abdominal pain | 1.0 | 0.47 | 0.30 | 1.0 | 0.57 |
| Shortness of breath | 0.91 | 0.59 | 0.33 | 0.97 | 0.64 |
| Confusion | 0.66 | 0.93 | 0.44 | 0.97 | 0.90 |
| Liver cirrhosis | 1.0 | 0.67 | 0.15 | 1.0 | 0.69 |

| | | | | | |
|---|---|---|---|---|---|
| Ascites | 1.0 | 0.51 | 0.07 | 1.0 | 0.53 |
| Abdominal pain | 0.97 | 0.41 | 0.27 | 0.98 | 0.51 |
| Shortness of breath | 0.92 | 0.59 | 0.33 | 0.97 | 0.65 |
| Confusion | 0.62 | 0.87 | 0.30 | 0.96 | 0.85 |
| Liver cirrhosis | 1.0 | 0.83 | 0.24 | 1.0 | 0.84 |

| | | | | | |
|---|---|---|---|---|---|
| Ascites | 0.95 | 0.94 | 0.38 | 1.00 | 0.94 |
| Abdominal pain | 0.99 | 0.18 | 0.22 | 0.99 | 0.33 |
| Shortness of breath | 0.64 | 0.95 | 0.72 | 0.92 | 0.89 |
| Confusion | 0.71 | 0.85 | 0.30 | 0.97 | 0.84 |
| Liver cirrhosis | 1.00 | 0.65 | 0.14 | 1.00 | 0.67 |

| | | | | | |
|---|---|---|---|---|---|
| Ascites | 1.0 | 0.57 | 0.09 | 1.0 | 0.59 |
| Abdominal pain | 0.98 | 0.53 | 0.32 | 0.99 | 0.62 |
| Shortness of breath | 0.91 | 0.67 | 0.37 | 0.97 | 0.71 |
| Confusion | 0.63 | 0.83 | 0.25 | 0.96 | 0.82 |
| Liver cirrhosis | 1.0 | 0.02 | 0.05 | 1.0 | 0.07 |

**Supplementary Figure 2 - Llama-2 Prompt engineering: Integration of System and User Prompts with 7b Model Evaluation.** This illustrates the Llama-2 prompt engineering process, highlighting two distinct modules: the system prompt and the user prompt. The system prompt is designed to guide the behavior of the Language Model, setting the overall context and parameters for interaction. Following this, the user prompt provides detailed, specific instructions and questions, tailoring the model's response to particular tasks or queries. In the first round of prompt engineering with the 7b model, both the system and user prompt were employed. (1) We included a report and the respective questions about the features in the user prompt and then compared the accuracy of this prompting technique with the following approaches, which include more modules: (2) Adapting a chain-of-thought (CoT) approach, where the model was prompted to provide the corresponding text excerpt before answering the questions and grammar-forced output of this excerpt did not enhance the accuracy. (3) Giving an example deteriorated the accuracy for all features except for shortness of breath. (4) Providing an additional definition about the features improved the accuracy for the features shortness of breath and abdominal pain. Npv= Negative predictive value, Ppv= Positive predictive value.

**7B** | **70B**

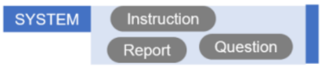


7B:

| Features | Sensitivity | Specificity | Ppv | Npv | Accuracy |
|---|---|---|---|---|---|
| Ascites | 1.0 | 0.81 | 0.18 | 1.0 | 0.82 |
| Abdominal pain | 0.98 | 0.55 | 0.33 | 0.99 | 0.63 |
| Shortness of breath | 0.92 | 0.63 | 0.35 | 0.97 | 0.68 |
| Confusion | 0.66 | 0.95 | 0.53 | 0.97 | 0.92 |
| Liver cirrhosis | 1.0 | 0.74 | 0.18 | 1.0 | 0.76 |

70B:

| Feature | Sensitivity | Specificity | Ppv | Npv | Accuracy |
|---|---|---|---|---|---|
| Ascites | 0.95 | 0.99 | 0.86 | 1.0 | 0.99 |
| Abdominal pain | 0.90 | 0.93 | 0.74 | 0.98 | 0.92 |
| Shortness of breath | 0.93 | 0.90 | 0.67 | 0.98 | 0.91 |
| Confusion | 0.83 | 0.93 | 0.52 | 0.98 | 0.92 |
| Liver cirrhosis | 1.0 | 0.96 | 0.59 | 1.0 | 0.96 |

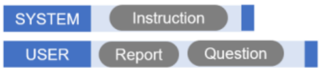


7B:

| Feature | Sensitivity | Specificity | Ppv | Npv | Accuracy |
|---|---|---|---|---|---|
| Ascites | 0.95 | 0.78 | 0.15 | 1.0 | 0.78 |
| Abdominal pain | 1.0 | 0.47 | 0.30 | 1.0 | 0.57 |
| Shortness of breath | 0.91 | 0.59 | 0.33 | 0.97 | 0.64 |
| Confusion | 0.66 | 0.93 | 0.44 | 0.97 | 0.90 |
| Liver cirrhosis | 1.0 | 0.67 | 0.15 | 1.0 | 0.69 |

70B:

| Feature | Sensitivity | Specificity | Ppv | Npv | Accuracy |
|---|---|---|---|---|---|
| Ascites | 1.0 | 0.9 | 0.29 | 1.0 | 0.90 |
| Abdominal pain | 0.91 | 0.91 | 0.71 | 0.98 | 0.91 |
| Shortness of breath | 0.92 | 0.92 | 0.73 | 0.98 | 0.92 |
| Confusion | 0.88 | 0.93 | 0.53 | 0.99 | 0.93 |
| Liver cirrhosis | 1.0 | 0.95 | 0.53 | 1.0 | 0.95 |

**Supplementary Figure 3 - Performance with all Modules in System and User Prompt.**

Npv= Negative predictive value, Ppv= Positive predictive value.

### Supplementary Methods

#### Prompt engineering strategy

We delved into the effectiveness of different prompt architectures. This involved a detailed exploration of the roles and impacts of system and user prompts. The system prompt was designed to provide initial instructions or explanations to guide the interaction, while the user prompt included the user's input or query, further shaping the response process. For explainability purposes, we forced the model via grammar to consistently output an excerpt from the text before answering the question. We identified and utilized several key prompt modules: General Instructions, Report, Questions, Chain-of-thought (CoT) Questions (where the model was specifically asked to provide an excerpt from text and then answer the question, see **Chain-of-thought Prompt**), Definitions (see **Definition Prompt)**, and Example (see **One-Shot Prompt)**. Each of these modules served a distinct purpose. General Instructions provided the basic guidelines for the interaction, the report module delivered specific the present medical history, the questions module consisted of queries to be addressed by the system, CoT aimed to elicit detailed, sequential responses, definitions offered explanations of terms or concepts, and the example module provided an illustration to guide response formation. Our methodological approach involved iterating various combinations of these modules within both the system and user prompts. The objective was to assess how different configurations affected the model's performance, providing a comprehensive understanding of the most effective prompt structures(Supplementary Figure 2 and 3). The MAIN ZERO SHOT PROMPT shows the final zero shot prompt, underlying the results in **Figure 3** and **Figure 5A.**

#### MAIN ZERO SHOT PROMPT

[INST] <<SYS>>

You are programmed as a cooperative medical assistant. A patient report will be available to you, and users will request specific information from this report. Your responses should adhere rigorously to the information contained within the provided report, ensuring no fabrication or assumption of details not explicitly stated

**This is the report:**

{}

**Now answer following questions:**

From the report, is ascites present at or before patient admission?

From the report, is abdominal pain present at or before admission?

From the report, is shortness of breath present at or before admission?

From the report, is confusion present at or before admission?

From the report, is liver cirrhosis present or suspected at admission?

**These are the definitions:**

**Abdominal pain** refers to any discomfort or pain that occurs in the abdominal area. It may sometimes be abbreviated as "abd pain" in medical contexts. The pain can also be specifically located and described by its region: Epigastric: Near the upper-middle region of the abdomen. RUQ: Right Upper Quadrant. RLQ: Right Lower Quadrant. LUQ: Left Upper Quadrant. LLQ: Left Lower Quadrant. If a 10-point review of systems (ROS) does not indicate any issues (is described as negative) and "abdominal pain" or its abbreviation are not explicitly mentioned in a medical report, it indicates that the patient does not have abdominal pain per the context provided in the definition.

**Shortness of breath** (also known as SOB or dyspnea) refers to difficulty breathing. If it occurs during physical activity, it's referred to as dyspnea on exertion (DOE). If a 10 point review of systems (ROS) is negative (i.e., does not indicate any abnormality or issue) and the terms "dyspnea," "SOB," or "DOE" are not otherwise mentioned in a medical report, this is taken to mean the patient is not experiencing shortness of breath according to the context given.

**Confusion** is a mental state characterized by disorientation and an inability to think clearly, often manifesting as difficulty remembering, making decisions, and maintaining awareness of critical aspects such as time, place, and personal identity. In medical contexts, the concept of orientation is pivotal. 'Oriented x4' indicates that an individual is lucid and aware of four key domains: person (awareness of oneself), place (recognition of physical location), time (understanding of the day, date, and/or time), and situation (comprehension of the ongoing events or circumstances). Consequently, being 'Oriented x4' signifies the absence of confusion. Conversely, if orientation is noted as less than 4, e.g., 'oriented x3', confusion is presumed present. Furthermore, impaired vigilance, exemplified when a patient is only intermittently responsive, is also indicative of confusion. Practical examples from medical reports might include phrases such as 'pt has brief period of confusion' or 'alert-oriented x3', suggesting episodes or states of confusion within the patient's condition.

<</SYS>>

[/INST]

#### One-Shot Prompt

[INST] <<SYS>>

You are programmed as a cooperative medical assistant. A patient report will be available to you, and users will request specific information from this report. Your responses should adhere rigorously to the information contained within the provided report, ensuring no fabrication or assumption of details not explicitly stated. You will be given an example, then proceed with the report provided.

<</SYS>> [/INST]

This is the report:

___ HCV cirrhosis c/b ascites, hiv on ART, h/o IVDU, COPD, \nbioplar, PTSD, presented from OSH ED with worsening abd \ndistension over past week. \nPt reports self-discontinuing lasix and spirnolactone ___ weeks \nago, because she feels like \"they don't do anything\" and that \nshe \"doesn't want to put more chemicals in her.\" She does not \nfollow Na-restricted diets. In the past week, she notes that she \nhas been having worsening abd distension and discomfort. She \ndenies ___ edema, or SOB, or orthopnea. She denies f/c/n/v, d/c, \ndysuria. She had food poisoning a week ago from eating stale \ncake (n/v 20 min after food ingestion), which resolved the same \nday. She denies other recent illness or sick contacts. She notes \nthat she has been noticing gum bleeding while brushing her teeth \nin recent weeks. she denies easy bruising, melena, BRBPR, \nhemetesis, hemoptysis, or hematuria. \nBecause of her abd pain, she went to OSH ED and was transferred \nto ___ for further care. Per ED report, pt has brief period of \nconfusion - she did not recall the ultrasound or bloodwork at \nosh. She denies recent drug use or alcohol use. She denies \nfeeling confused, but reports that she is forgetful at times. \nIn the ED, initial vitals were 98.4 70 106/63 16 97%RA \nLabs notable for ALT/AST/AP ___ ___: ___, \nTbili1.6, WBC 5K, platelet 77, INR 1.6

Now answer following questions:

From the report, is ascites present at or before patient admission?

From the report, is abdominal pain present at or before admission?

From the report, is shortness of breath present at or before admission?

From the report, is confusion present at or before admission?

From the report, is liver cirrhosis present or suspected at admission?

Example output:

{"content": "{\n\"ascites\":\n{\n\"excerpt\": \"HCV cirrhosis c/b ascites\"\n,\"present\": true\n}\n,\"abdominal pain\": {\n\"excerpt\": \"worsening abd distension over past week\"\n,\"present\": true\n}\n,\"shortness of breath\": {\n\"excerpt\": \"denies SOB\"\n,\"present\": false\n}\n,\"confusion\": {\n\"excerpt\": \"brief period of confusion - she did not recall the ultrasound or bloodwork at osh\"\n,\"present\": true\n}\n,\"liver cirrhosis\": {\n\"excerpt\": \"HCV cirrhosis c/b ascites\"\n,\"present\": true\n}\n}\n"}

[INST]

This is the report:

{{REPORT}}

Now answer following questions:

From the report, is ascites present at or before patient admission?

From the report, is abdominal pain present at or before admission?

From the report, is shortness of breath present at or before admission?

From the report, is confusion present at or before admission?

From the report, is liver cirrhosis present or suspected at admission?

[/INST]"""

#### Chain-of-thought Prompt

[INST] <<SYS>>

You are programmed as a cooperative medical assistant. A patient report will be available to you, and users will request specific information from this report. Your responses should adhere rigorously to the information contained within the provided report, ensuring no fabrication or assumption of details not explicitly stated. Provide an excerpt from text first, then answer the questions.

<</SYS>>

Please extract the following information from the text:

Is **ascites** present at admission? Provide an excerpt from text, then answer the question.

Is **abdominal pain** present at or before admission? Provide an excerpt from the text, then answer the question.

Is **shortness of breath** present at or before admission? Provide an excerpt from the text, then answer the question.

Is **confusion** present at or before admission? Provide an excerpt from the text, then answer the question.

Is **liver cirrhosis** present or suspected at admission? Provide an excerpt from the text, then answer the question.

[/INST]

#### Definition Prompt

[INST] <<SYS>>

You are programmed as a cooperative medical assistant. A patient report will be available to you, and users will request specific information from this report. Your responses should adhere rigorously to the information contained within the provided report, ensuring no fabrication or assumption of details not explicitly stated.

<</SYS>>

**This is the report:**

{}

Now answer following questions:

From the report, is ascites present at or before patient admission?

From the report, is abdominal pain present at or before admission?

From the report, is shortness of breath present at or before admission?

From the report, is confusion present at or before admission?

From the report, is liver cirrhosis present or suspected at admission?

**These are the definitions:**

**Ascites** refers to the accumulation of excess fluid in the peritoneal cavity, which is the space between the organs and the abdominal wall, often resulting from liver disease, heart failure, or cancer.

**Abdominal pain** refers to any discomfort or pain that occurs in the abdominal area. It may sometimes be abbreviated as "abd pain" in medical contexts. The pain can also be specifically located and described by its region: Epigastric: Near the upper-middle region of the abdomen. RUQ: Right Upper Quadrant. RLQ: Right Lower Quadrant. LUQ: Left Upper Quadrant. LLQ: Left Lower Quadrant. If a 10-point review of systems (ROS) does not indicate any issues (is described as negative) and "abdominal pain" or its abbreviation are not explicitly mentioned in a medical report, it indicates that the patient does not have abdominal pain per the context provided in the definition.

**Shortness of breath**: Shortness of breath (also known as SOB or dyspnea) refers to difficulty breathing. If it occurs during physical activity, it's referred to as dyspnea on exertion (DOE). If a 10 point review of systems (ROS) is negative (i.e., does not indicate any abnormality or issue) and the terms "dyspnea," "SOB," or "DOE" are not otherwise mentioned in a medical report, this is taken to mean the patient is not experiencing shortness of breath according to the context given.

**Confusion** is a mental state characterized by disorientation and an inability to think clearly, often manifesting as difficulty remembering, making decisions, and maintaining awareness of critical aspects such as time, place, and personal identity. In medical contexts, the concept of orientation is pivotal. 'Oriented x4' indicates that an individual is lucid and aware of four key domains: person (awareness of oneself), place (recognition of physical location), time (understanding of the day, date, and/or time), and situation (comprehension of the ongoing events or circumstances). Consequently, being 'Oriented x4' signifies the absence of confusion. Conversely, if orientation is noted as less than 4, e.g., 'oriented x3', confusion is presumed present. Furthermore, impaired vigilance, exemplified when a patient is only intermittently responsive, is also indicative of confusion. Practical examples from medical reports might include phrases such as 'pt has brief period of confusion' or 'alert-oriented x3', suggesting episodes or states of confusion within the patient's condition.

**Liver cirrhosis:** Is a late stage of scarring (fibrosis) of the liver caused by many forms of liver diseases and conditions, such as hepatitis and chronic alcoholism, leading to loss of liver function and potential complications like bleeding, jaundice, and hepatic encephalopathy. Examples:

HCV cirrhosis, decompensated alcoholic and Hepatitis C cirrhosis, ETO cirrhosis.

[/INST]""""
